# Zombie trials are often monocentric and cluster within fabricated evidence “factories”: a cohort study of 236 retracted randomized controlled trials with confirmed data fabrication

**DOI:** 10.64898/2026.07.23.26357975

**Authors:** Tala Jajieh, Céline Chapelle, Cédric Lemarchand, Clara Locher, Marc-Antoine Pencolé, John P.A. Ioannids, Edouard Ollier, Florian Naudet, Silvy Laporte

**Affiliations:** Université Jean-Monnet, Mines Saint-Étienne, INSERM Unité 1059, Santé Ingénierie Biologie Saint-Étienne (SAINBIOSE), Saint-Étienne, France; Service de Pharmacologie Clinique, CHU de Saint-Étienne, France; Univ Rennes, Inserm, EHESP, Irset (Institut de recherche en santé, environnement et travail) – UMR_S 1085, F-35000 Rennes, France; Université Paris Cité, Sorbonne Université, Inserm, Centre de Recherche des Cordeliers, F-75006 Paris, France; Stanford Prevention Research Center, Department of Medicine, Stanford University School of Medicine, Stanford, California, USA; Department of Epidemiology and Population Health, Stanford University School of Medicine, Stanford, California, USA; Department of Biomedical Data Science, Stanford University School of Medicine, Stanford, California, USA; Meta-Research Innovation Center at Stanford (METRICS), Stanford University, Stanford, California, USA; University of Rennes CHU Rennes, Inserm, EHESP, IRSET (Institut de recherche en santé, environnement et travail), Rennes, France; Institut Universitaire de France (IUF), Paris, France

**Keywords:** zombie trials, data fabrication, retraction, research integrity, scientific misconduct, evidence-based medicine

## Abstract

**Background:** Randomized controlled trials (RCTs) that rely on fabricated or fictitious data, commonly known as “zombie” trials, threaten evidence-based medicine by contaminating systematic reviews and clinical guidelines. We describe a cohort of zombie trials with confirmed data fabrication.

**Methods:** The cohort was derived from the Retraction Watch database by updating and extending the dataset used in the VITALITY study to include all retracted zombie trials identified from database inception through December 11, 2025. To ensure that included RCTs were identified as zombie trials rather than retracted for other reasons, retraction notices were text-mined for terms indicative of data fabrication. Trial- and author-level characteristics were extracted using a combination of automated and manual methods by one reviewer. We applied descriptive statistics, co-authorship network analysis and cascade effect mapping.

**Results:** Among 236 retracted RCTs published between 1983 and 2024 and identified as zombie trials with confirmed data fabrication, 197 (83.5%) were associated with authors who had a history of repeated misconduct. These trials involved 450 authors overall, including only 66 unique first authors, with one author alone accounting for 104 trials (43.8% of the cohort). Authors associated with multiple zombie trials experienced substantially longer delays to retraction, with a median of 13.6 years (IQR 8.0–15.3), compared with 2.75 years for authors linked to a single zombie trial. Most retracted zombie RCTs were monocentric (91.9%), involved a median of 3.5 authors per trial, and predominantly evaluated pharmacological interventions. They were largely concentrated in anesthesiology (58.1%) and in Japan (56.4%). Adjusted for the volume of trials produced by country, Japan, Tunisia, and Egypt had the highest rates. Co-authorship networks formed fragmented, largely disconnected clusters, suggesting localized “factories” of fabricated evidence. Retraction cascades were common, with the identification of a single fraudulent trial often leading to chains of retractions that exposed misconduct spanning decades.

**Conclusion:** Confirmed zombie trials were usually generated from single centers that published many of such trials.

## Introduction

Scientific integrity is the cornerstone of reliable and credible research(1), but it is increasingly threatened by scientific misconduct, including data fabrication, falsification, and plagiarism (2,3). The number of retracted articles has risen sharply over recent decade and as of June 2026, the Retraction Watch database contained more than 65,000 retracted papers affected by serious integrity concerns (4). These numbers represent only “the tip of the iceberg” (5), as official retraction rates vastly underestimates the true prevalence of problematic research. Furthermore, due to the protracted nature of the retraction process, many flawed studies continue to circulate in the literature (6).

Because randomized controlled trials (RCTs) are considered one of the highest levels of evidence and underpin systematic reviews and meta-analyses of medical interventions (7,8). Research misconduct in this area poses a major threat to evidence-based medicine. Even a single fraudulent trial can contaminate evidence syntheses, distort clinical guidelines, and ultimately harm patients (9,10). When such misconduct is uncovered, it typically triggers a wave of retractions (11). To illustrate, the Japanese researcher Yoshihiro Sato fabricated data across dozens of trials (12), resulting in 113 retracted papers. His fraudulent work was cited before retraction in 88 systematic reviews and clinical guidelines, some of which directly informed Japan’s osteoporosis treatment recommendations (13). Trials with fabricated data have been referred to as “zombie trials”. Zombie trials have fundamental flaws in their data due to fabrication, falsification, unethical practices, or inadequate quality controls, rendering them scientifically invalid and untrustworthy (14,15). Hundreds of thousands of zombie trials may circulate within biomedical literature (14). Some highly speculative estimates have even suggested that between one-quarter and one-third of all RCTs may be fabricated or unreliable, with several medical specialties such as women’s health and COVID-19 ivermectin research being disproportionately affected (8).

It is therefore important to improve the detection of zombie trials (16), either by identifying potential red flags before publication or by facilitating their rapid detection and retraction post-publication, in order to prevent their dissemination. However, there is currently limited empirical evidence on the typical characteristics and systemic markers of zombie trials that could help raise early suspicion during the editorial process. A recent initiative is the development and endorsement by the Cochrane Collaboration of the INSPECT-SR (INveStigating ProblEmatic Clinical Trials in Systematic Reviews) tool (17), which helps systematic reviewers assess the trustworthiness of clinical trials and make informed decisions about whether they should be included in evidence syntheses. We therefore assembled a cohort of retracted zombie trials to support their identification and characterization, as well as the subsequent validation of INSPECT-SR. There is no gold standard for identifying zombie trials even when they are formally retracted, because retraction notices are often vague or insufficiently informative. We therefore restricted our cohort to retracted studies for which data fabrication was explicitly documented. This report describes the cohort construction and the descriptive characteristics of the included trials.

## Methods

### Protocol and study registration

The protocol was prospectively registered on the Open Science Framework (OSF) (https://doi.org/10.17605/OSF.IO/NGEVC), and reporting followed the PRISMA guideline (18,19).

### Design and selection criteria

To build the cohort of confirmed zombie trials, a two-step selection process was implemented. First, we considered all retracted RCTs involving human participants who evaluated the effects of medical interventions to be eligible, regardless of study design (parallel, crossover, cluster, open-label or blind), medical field, pathology, or intervention type. Then, to ensure that included RCTs qualified as zombie trials rather than being retracted for other reasons, retraction notices were text-mined for terms specifically indicative of data fabrication.

### Source

We anchored our source data on the VITALITY cohort of RCTs identified by Xu et al. (10) via the Retraction Watch database (20) from inception to November 5, 2024, with no restrictions on language or publication period. This parent cohort is openly available (https://doi.org/10.17605/OSF.IO/NGEVC). From this subset of 1330 retracted trials, trials formally retracted due to data fabrication were included, whereas those retracted solely due to concerns related to data integrity or errors were excluded. The search strategy by Xu et al. was reviewed by two information specialists in the original study. We updated this search on December 11, 2025, adhering strictly to the original methodology. This involved executing “two separate searches to retrieve records labelled as ‘Clinical Study’ and ‘Research Article’, while restricting the scope to the Health Sciences (HSC) subject domain” using the Retraction Watch Database.

### Selection process of retracted zombie trials

Following the updated search, we systematically screened the records to identify retracted zombie trials. This identification process relied on the triangulation of three distinct data sources: first, the standardized reasons for retraction automatically indexed within the Retraction Watch database, which specifies why an article was retracted using specific terms; second, the full text of original retraction notices extracted manually from PubMed, which contain formal statements from authors, journal editors, or publisher explaining the reason for the retraction of the article; and third, post-publication peer reviews and public comments extracted manually from the PubPeer platform (21). Based on a preliminary review of these sources, a specific set of semantic keywords characterizing data fabrication (e.g., ‘fictive data’, ‘fabricated data’, ‘resulting from imagination’) was defined. The complete list of keywords is provided in Appendix I. Text mining techniques were then executed using R Software to parse the retraction text fields and programmatically isolate trials matching these definitive keywords.

### Data extraction

A dedicated database of retracted zombie trials was built, initially containing 24 variables derived from the original dataset by Xu et al. (10). These basic metadata included the DOI, article title, journal name, retraction reasons, retraction notes, paywall status, involvement of paper mills, total citation counts, retraction initiator, publication and retraction dates, funding sources, and trial registry information. To extensively characterize these trials, this dataset was expanded through a combination of automated and manual data extraction workflows.

Automated extraction was performed using PubMed and Retraction Watch databases to retrieve additional characteristics, including country of the first author and all authors, publisher, medical specialty, recommendable journal status (possible non-predatory), Journal, year of publication, retraction date, and others. As PubMed does not provide a standardized publisher variable, journal titles were first mapped to their corresponding publishers using the Crossref API (22). Journals were grouped by publisher, and the number of PubMed-indexed articles associated with each publisher was estimated by querying PubMed through the Entrez API using journal titles. These publication counts were then used as the reference denominator to compare the distribution of publishers between retracted zombie trials and baseline published articles.

Manual extraction was performed to capture additional publication-related characteristics, including sample size, p-value associated with the primary endpoint, specific types of intervention, first author identity, and author approval for retraction. A complete list of these finalized extracted variables is provided in Appendix II, with the full methodological details provided in the study protocol (https://doi.org/10.17605/OSF.IO/NGEVC). One reviewer manually extracted author-level data for the first and last authors using Research Gate, LinkedIn, Google Scholar, and PubMed. These data included sex, /institutional affiliation, clinical specialty, and academic credentials (MD vs non-MD status). To minimize the risk of misattribution due to author name homonyms, author identity across records was validated using institutional affiliation, clinical specialty, and other identifying biographical information available on these platforms, ensuring that retraction histories were attributed only when sufficient concordance confirmed a shared identity. To evaluate the track record of misconduct, we transformed our updated full dataset (VITALITY) from a trial-level to an author-level structure, to automatically calculate the cumulative number of all retracted trials for each unique first and last author, as well as the number of non-zombie retracted trials, obtained by subtracting the number of zombie trials from the total number of retracted trials for each author. Within this framework, authors were classified as having multiple zombie retractions (≥2 independent trials) or a single zombie retraction (one trial) within the cohort.

### Statistical analysis

Descriptive statistics were used to characterize the retracted zombie trials cohort. Two distinct statistical units of analysis were considered: 1) the retracted zombie publications and 2) the individual authors involved in these publications. Categorical variables were summarized using numbers and percentages, while continuous variables were expressed as medians with interquartile ranges (IQRs, the difference between Q3 and Q1). Geospatial mapping was performed to illustrate the worldwide distribution of retracted zombie trials according to the first author’s country of affiliation and the geographic distribution of all authors according to their affiliation countries. To account for differences in national clinical trial output, country-level counts of retracted zombie trials, based on the first author’s country of affiliation, were further expressed as a ratio to the total number of registered clinical trials per country, obtained from the WHO Global Observatory on Health Research and Development. (23)

Plots were used to identify the most prolific authors involved in these retractions. The delay to retraction was calculated as the exact time elapsed between the initial publication date and the formal retraction date. To evaluate collaborative relationships, a co-authorship network was constructed where nodes represented unique authors and edges (connecting lines) represented co-authorship links. The distribution of retracted zombie trials across publishers was evaluated relative to their overall publication volume indexed in PubMed.

Furthermore, concentration indices of “retracted zombie trials” were estimated to quantify the distribution asymmetry of retracted zombie publications among the author pool, measuring the extent to which fraud is concentrated within a small group of individuals.

All statistical analyses and visualizations were performed using R software (version 4.4.1) and RStudio version 2026.05.0+218 (Golden Wattle).

### Changes to the initial protocol

Several amendments were made to the original protocol. As the work progressed, a comparative analysis between zombie retracted trials with authors having single versus multiple retracted zombie trials was added. This comparison was not initially planned but emerged as a natural and meaningful extension of our primary objectives, shedding light on what distinguishes retracted zombie trials from the broader literature in terms of both publication characteristics and the profile of first and last authors. Finally, validation of INSPECT-SR, as prespecified in the protocol, is ongoing and will be reported separately.

## Results

### Study selection

Our updated search for the VITALITY cohort of Xu et al.(10) identified 843 supplementary articles across the two databases (Figure 1). After removing 13 duplicates, screening 830 titles and abstracts, 763 records were excluded. This process yielded 67 additional retracted RCTs, bringing the original cohort of 1,330 up to a total of 1,397 retracted RCTs meeting the inclusion criteria. Following a comprehensive analysis of the retraction reasons and the application of text-mining techniques to this final pool of 1,397 retracted RCTs, we identified a total of 236 retracted zombie trials (16.9%), which formed our definitive study cohort. The remaining 1,161 retracted RCTs were excluded because their retraction reasons did not meet our text-mining criteria for confirmed data fabrication (Figure 1).

**Figure 1:**
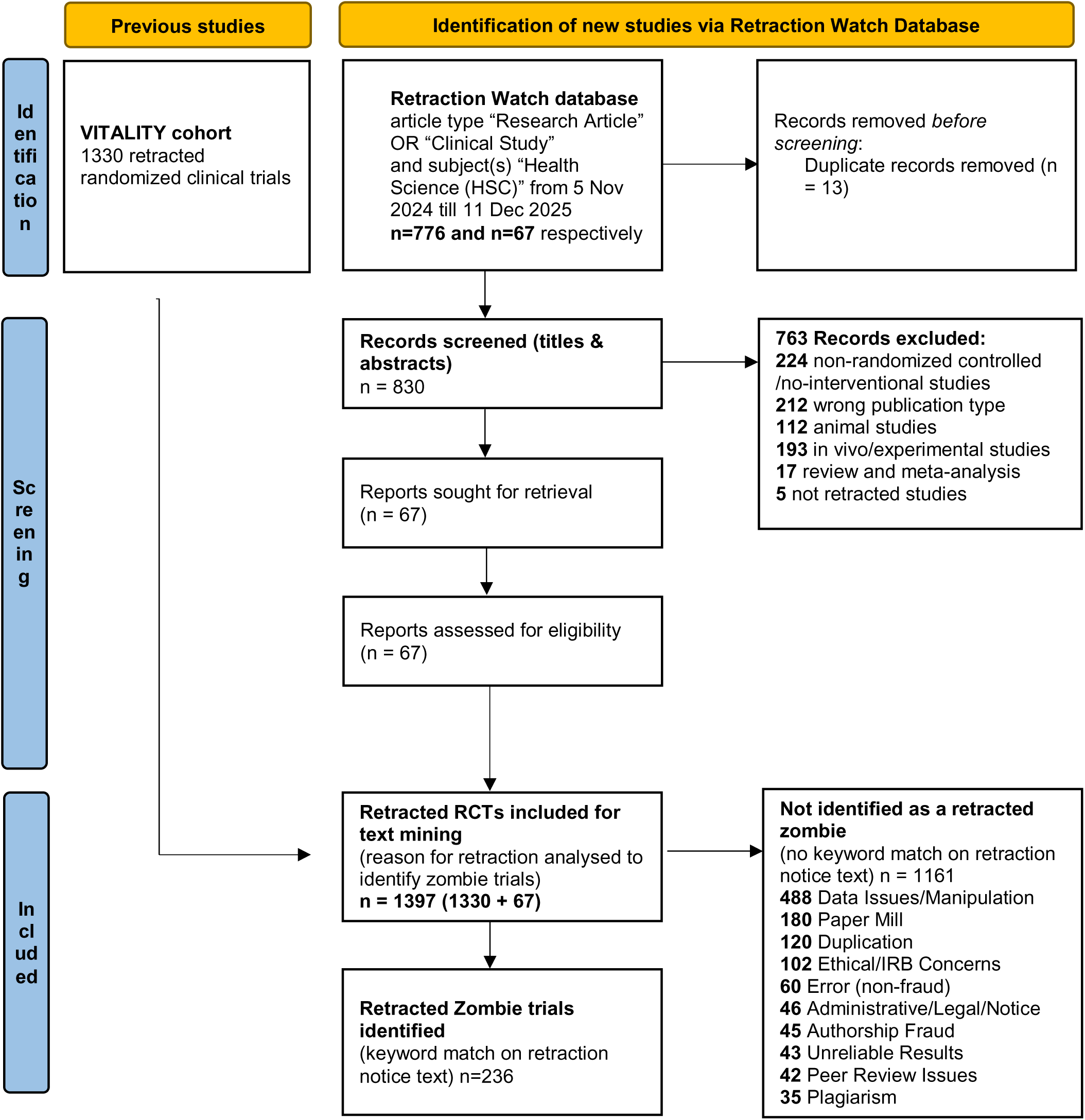
Flowchart of selection of retracted zombie trials

### Characteristics of the 236 retracted zombie trials

Table 1 presents the characteristics of the retracted zombie trial, and according to the authors associated with one versus multiple retracted zombie trials. Of the 236 identified retracted zombie papers, 197 (83.5%) were associated with authors who had multiple retractions. Trials involving authors associated with multiple zombie trials had substantially longer delays to retraction, with a median of 13.6 years (IQR 8.0–15.3), compared with 2.75 years (IQR 1.1–5.6) for trials whose authors were associated with a single zombie trial. 66.5% of trials linked to authors associated with multiple retracted zombie trials were more likely to have been published before 2005, whereas 82.1% of trials associated with a single retracted zombie trial were more likely to have been published in or after 2005. Among trials published in 2005 or later, registration remained less frequently reported among trials linked to authors associated with multiple retracted zombie trials compared with those linked to authors associated with a single retracted zombie trial (12/66, 18.2% vs. 15/32, 46.9%). Trials linked to authors associated with multiple retracted zombie trials also more often failed to report funding sources (81.7% vs. 48.7%). The distribution of JIFs (≥2) was similar between groups (43.2% vs. 38.5%). However, trials associated with authors of a single retracted zombie trial were more frequently published in possible predatory (not recommendable) journals than those associated with authors of multiple retracted zombie trials (48.7% vs. 20.3%). In both groups, retractions were primarily initiated by journal editors, and author non-response was the most common pattern regarding retraction approval.

**Table 1:**
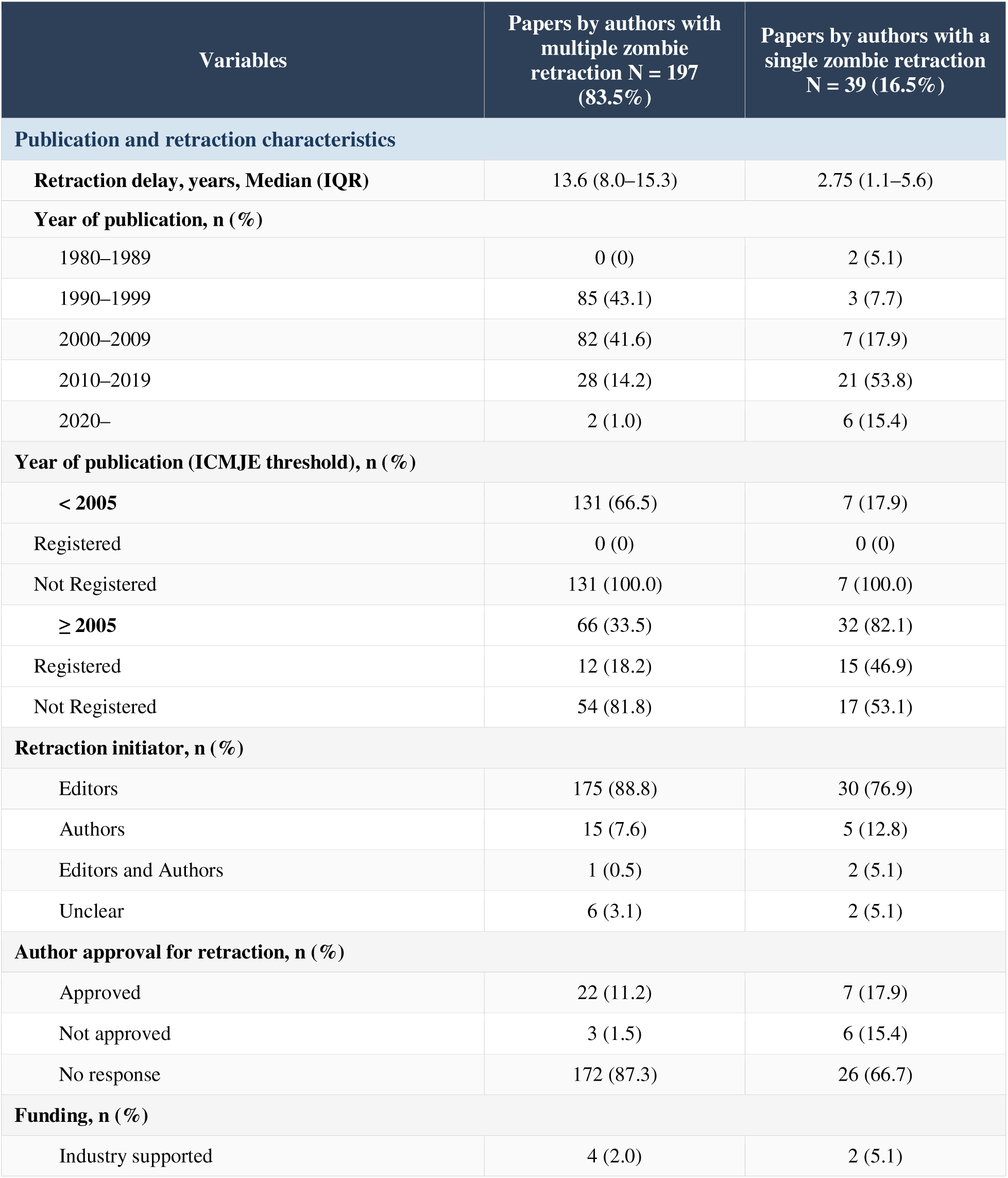
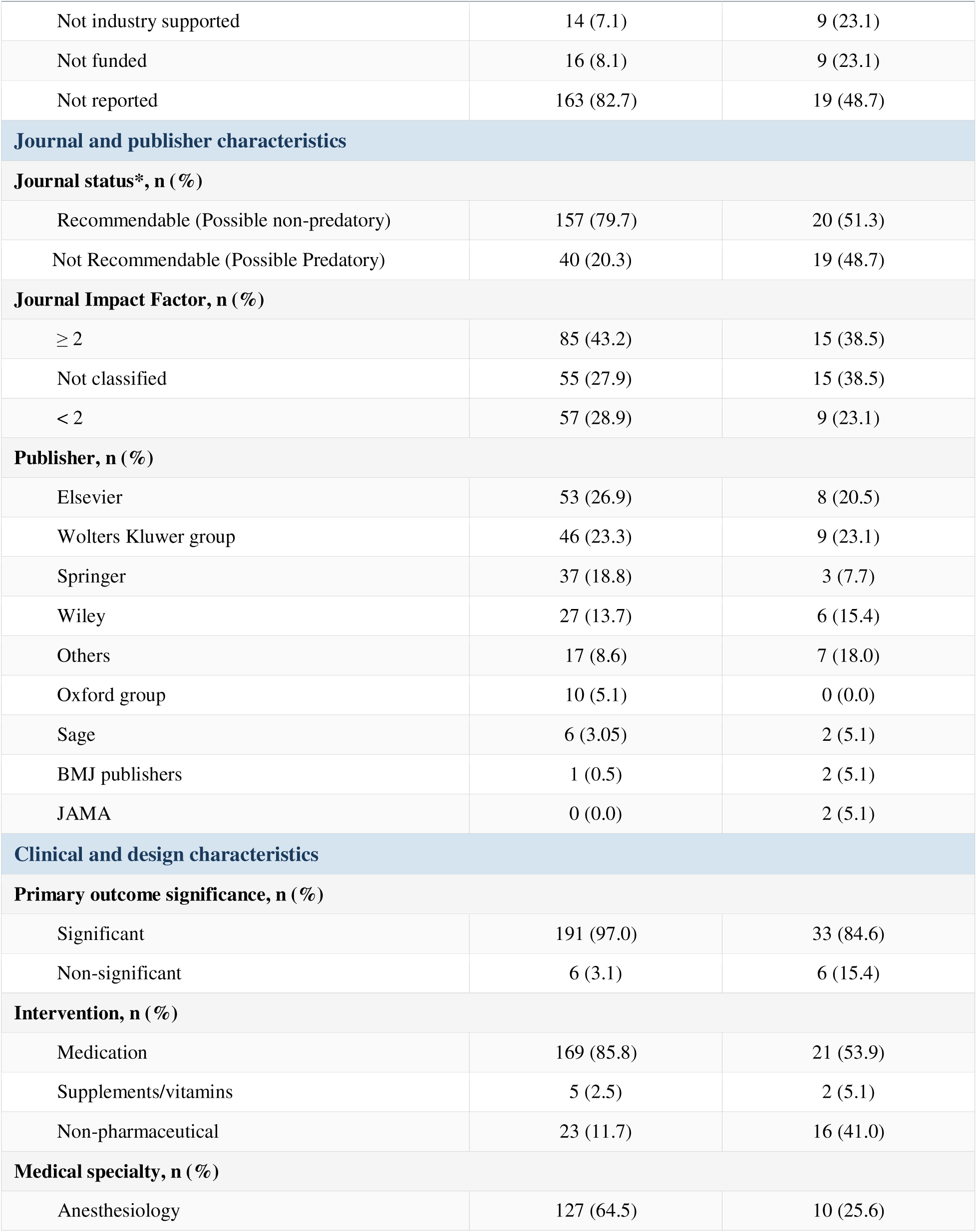
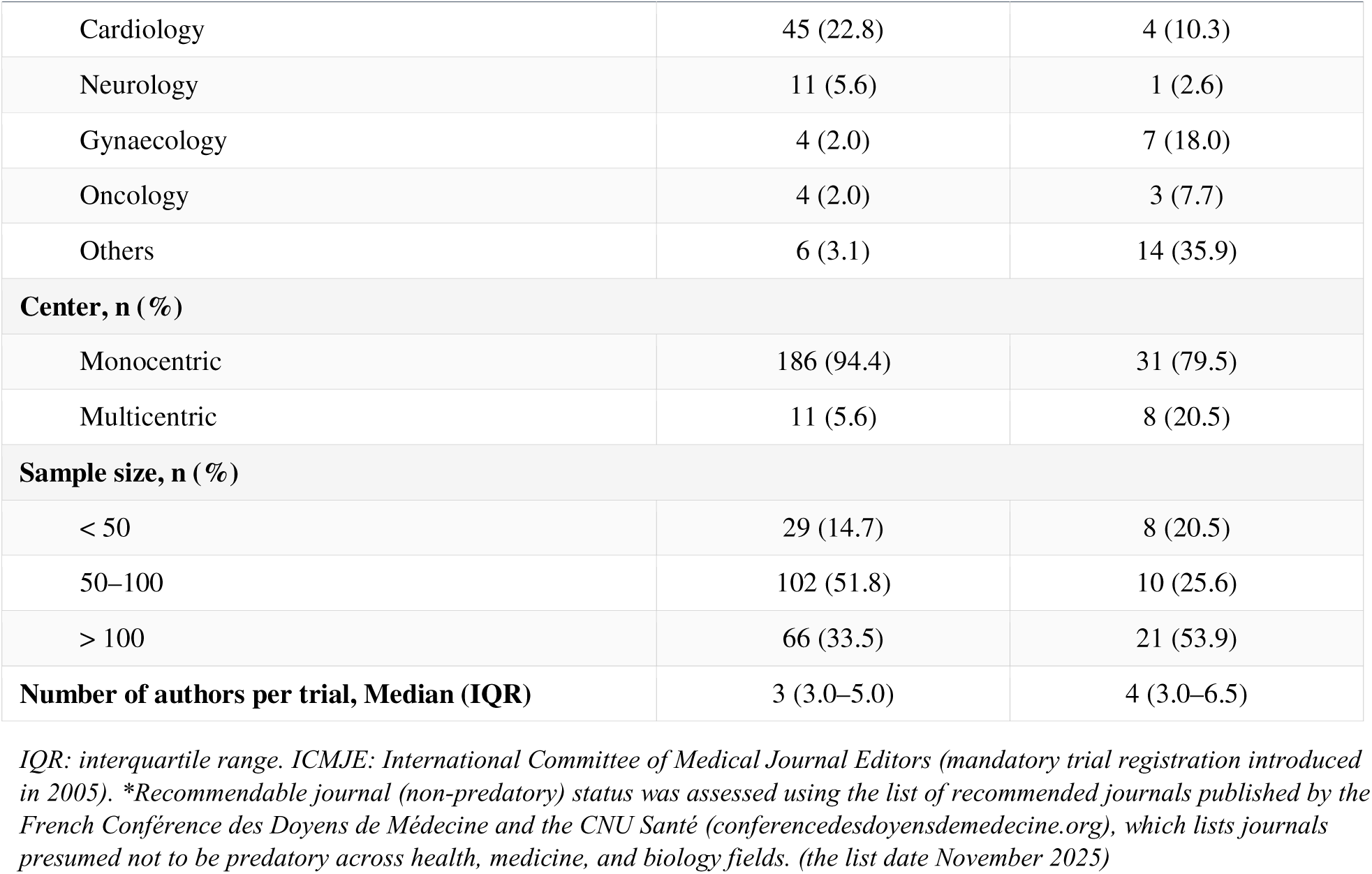
Characteristics of the 236 retracted zombie trials according to authors associated with one versus multiple retracted zombie trials.

| Variables | Papers by authors with multiple zombie retraction N = 197 (83.5%) | Papers by authors with a single zombie retraction N = 39 (16.5%) |
| --- | --- | --- |
| **Publication and retraction characteristics** |  |  |
| **Retraction delay, years, Median (IQR)** | 13.6 (8.0–15.3) | 2.75 (1.1–5.6) |
| **Year of publication, n (%)** |  |  |
| 1980–1989 | 0 (0) | 2 (5.1) |
| 1990–1999 | 85 (43.1) | 3 (7.7) |
| 2000–2009 | 82 (41.6) | 7 (17.9) |
| 2010–2019 | 28 (14.2) | 21 (53.8) |
| 2020– | 2 (1.0) | 6 (15.4) |
| **Year of publication (ICMJE threshold), n (%)** |  |  |
| < 2005 | 131 (66.5) | 7 (17.9) |
| Registered | 0 (0) | 0 (0) |
| Not Registered | 131 (100.0) | 7 (100.0) |
| ≥ 2005 | 66 (33.5) | 32 (82.1) |
| Registered | 12 (18.2) | 15 (46.9) |
| Not Registered | 54 (81.8) | 17 (53.1) |
| **Retraction initiator, n (%)** |  |  |
| Editors | 175 (88.8) | 30 (76.9) |
| Authors | 15 (7.6) | 5 (12.8) |
| Editors and Authors | 1 (0.5) | 2 (5.1) |
| Unclear | 6 (3.1) | 2 (5.1) |
| **Author approval for retraction, n (%)** |  |  |
| Approved | 22 (11.2) | 7 (17.9) |
| Not approved | 3 (1.5) | 6 (15.4) |
| No response | 172 (87.3) | 26 (66.7) |
| **Funding, n (%)** |  |  |
| Industry supported | 4 (2.0) | 2 (5.1) |
| Not industry supported | 14 (7.1) | 9 (23.1) |
| Not funded | 16 (8.1) | 9 (23.1) |
| Not reported | 163 (82.7) | 19 (48.7) |
| **Journal and publisher characteristics** |  |  |
| **Journal status\*, n (%)** |  |  |
| Recommendable (Possible non-predatory) | 157 (79.7) | 20 (51.3) |
| Not Recommendable (Possible Predatory) | 40 (20.3) | 19 (48.7) |
| **Journal Impact Factor, n (%)** |  |  |
| ≥ 2 | 85 (43.2) | 15 (38.5) |
| Not classified | 55 (27.9) | 15 (38.5) |
| < 2 | 57 (28.9) | 9 (23.1) |
| **Publisher, n (%)** |  |  |
| Elsevier | 53 (26.9) | 8 (20.5) |
| Wolters Kluwer group | 46 (23.3) | 9 (23.1) |
| Springer | 37 (18.8) | 3 (7.7) |
| Wiley | 27 (13.7) | 6 (15.4) |
| Others | 17 (8.6) | 7 (18.0) |
| Oxford group | 10 (5.1) | 0 (0.0) |
| Sage | 6 (3.05) | 2 (5.1) |
| BMJ publishers | 1 (0.5) | 2 (5.1) |
| JAMA | 0 (0.0) | 2 (5.1) |
| **Clinical and design characteristics** |  |  |
| **Primary outcome significance, n (%)** |  |  |
| Significant | 191 (97.0) | 33 (84.6) |
| Non-significant | 6 (3.1) | 6 (15.4) |
| **Intervention, n (%)** |  |  |
| Medication | 169 (85.8) | 21 (53.9) |
| Supplements/vitamins | 5 (2.5) | 2 (5.1) |
| Non-pharmaceutical | 23 (11.7) | 16 (41.0) |
| **Medical specialty, n (%)** |  |  |
| Anesthesiology | 127 (64.5) | 10 (25.6) |
| Cardiology | 45 (22.8) | 4 (10.3) |
| Neurology | 11 (5.6) | 1 (2.6) |
| Gynaecology | 4 (2.0) | 7 (18.0) |
| Oncology | 4 (2.0) | 3 (7.7) |
| Others | 6 (3.1) | 14 (35.9) |
| **Center, n (%)** |  |  |
| Monocentric | 186 (94.4) | 31 (79.5) |
| Multicentric | 11 (5.6) | 8 (20.5) |
| **Sample size, n (%)** |  |  |
| < 50 | 29 (14.7) | 8 (20.5) |
| 50–100 | 102 (51.8) | 10 (25.6) |
| > 100 | 66 (33.5) | 21 (53.9) |
| **Number of authors per trial, Median (IQR)** | 3 (3.0–5.0) | 4 (3.0–6.5) |

*IQR: interquartile range. ICMJE: International Committee of Medical Journal Editors (mandatory trial registration introduced in 2005). \*Recommendable journal (non-predatory) status was assessed using the list of recommended journals published by the French Conférence des Doyens de Médecine and the CNU Santé (conferencedesdoyensdemedecine.org), which lists journals presumed not to be predatory across health, medicine, and biology fields. (the list date November 2025)*

A statistically significant primary outcome was reported in 97.0% of trials associated with authors of multiple retracted zombie trials, compared with 83.6% of those associated with a single retracted zombie trial. Trials associated with authors of multiple retracted zombie trials predominantly evaluated pharmacological interventions (85.8% vs. 53.9%) and were largely concentrated in anesthesiology (64.5%), whereas trials associated with authors of a single retracted zombie trial were distributed across a broader range of medical specialties. Structurally, trials associated with authors of multiple retracted zombie trials were almost exclusively monocentric (94.4% vs. 79.5%).

These findings were corroborated by the multiple correspondence analysis (Appendix III), which showed the same distinction between trials with a single zombie retraction and trials linked to authors with multiple zombie retractions.

### Geographical distribution and publisher-level patterns

Comparing the raw number of retracted zombie trials per country to the ratio adjusted for each country’s total registered clinical trial volume (WHO Global Observatory on Health Research and Development) showed a shift in country ranking (Figure 2). After adjustment for national trial volume, the five countries with the highest ratios were Japan (1.8 per 1,000 trials), Tunisia (0.967), Egypt (0.868).

**Figure 2:**
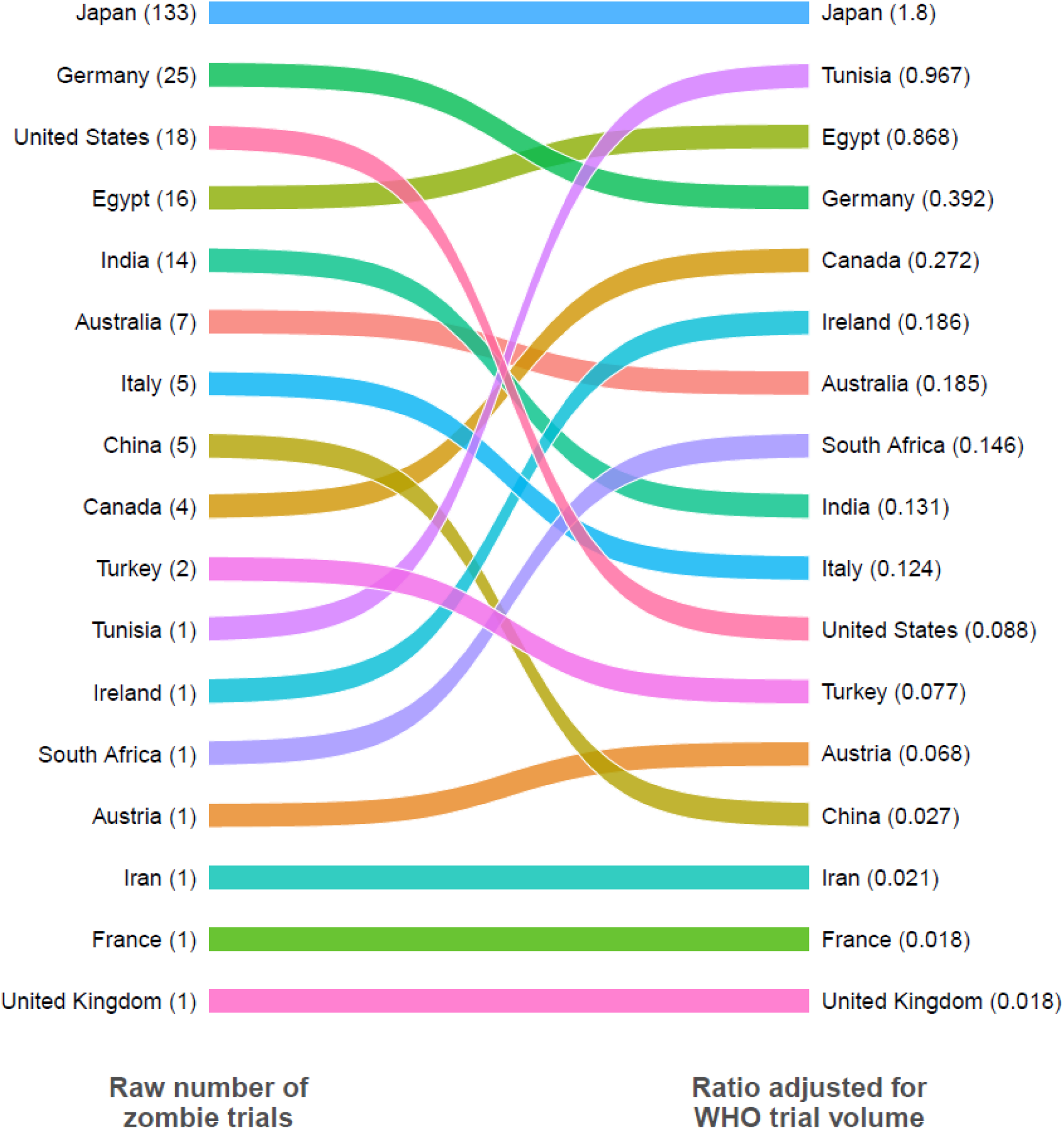
Shift in country ranking of retracted zombie trials after adjustment for national clinical trial volume. Comparison of country ranking by raw number of retracted zombie trials (left, per 236 total zombie trials) versus ranking adjusted for each country’s total registered clinical trial volume (right, per 1,000 registered clinical trials). Raw counts represent the number of retracted zombie trials with a first author affiliated to that country. Adjusted ratios represent the number of retracted zombie trials per 1,000 clinical trials registered in that country, using trial volume data from the WHO Global Observatory on Health Research and Development. Colored bands connect each country’s position across the two rankings; countries are ordered from highest to lowest within each column.

As a sensitivity analysis, we repeated the WHO-adjusted ranking using fractional country attribution, in which each trial’s contribution was divided equally among all countries represented among its authors, rather than the first author’s country alone (Appendix IV). Under this approach, Japan remained the highest-ranked country (1.81 per 1,000 trials), followed by Tunisia (0.967) and Egypt (0.841).

A world map showing the geographical distribution of all co-authors involved in the zombie trials, as opposed to first authors only, is provided in the (Appendix V). The distribution of first authors was concentrated in a smaller number of countries, whereas the distribution of all co-authors extended across a broader set of countries, including the United States, Germany, India, China, Australia, and several countries in the Middle East and Africa.

Variations were also observed across publishers (Appendix VI): The Wolters Kluwer Group accounted for 24% of retracted zombie trials compared to 6% of baseline published articles, whereas Taylor & Francis accounted for 16% of published articles but only 1% of the zombie trial cohort.

### Co-authorship Network Analysis

The 236 retracted zombie trials involved 450 unique authors, including 66 first authors and 100 last authors. The median number of authors per article was 3, with 163 papers (69.1%) featuring fewer than 5 authors. The co-authorship network of retracted zombie trials demonstrated no single dominant collaboration pattern (Figure 3). Instead, the network was highly fragmented into 51 isolated clusters ranging in size from 1 to 49 co-authors. 39 of these components represented isolated, single-trial author lists. In contrast, the remaining 12 clusters represented persistent, multi-paper research networks where specific subsets of co-authors repeatedly cross-collaborated. Within these, the largest collaborative group comprised 49 interconnected authors spanning 21 distinct retracted trials, anchored by Joachim Boldt. Analysis of edge weights further revealed localized, high-frequency partnerships, most notably a dominant pair (Yoshitaka Fujii and Hiroyoshi Tanaka) who exhibited an extraordinary co-authorship tie of 78 shared papers, represented by the darkest and thickest edges in the network layout.

**Figure 3:**
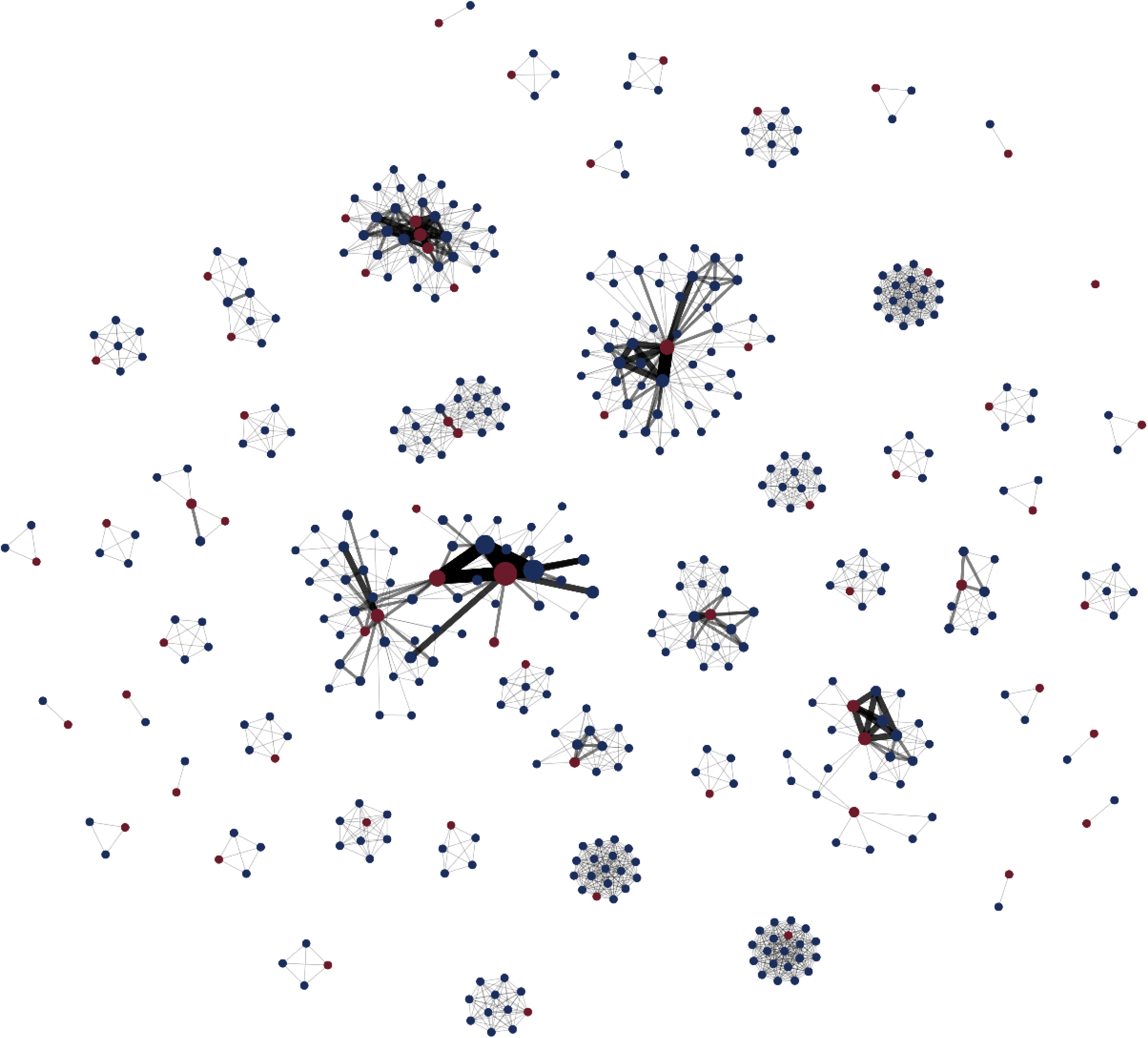
Co-Authorship Network diagram of retracted zombie trials’ authors. This network illustrates the relationships between 450 authors responsible for the 236 retracted zombie trials. Each node represents one author, and the connections (lines) between them indicate the co-authorship. Node size: the bigger the node, the greater the number of retracted zombie trials they were involved in. Node color: burgundy (red) dots are first authors, while navy blue dots represent co-authors. Edge (line) thickness and darkness: the more the edges are thick and dark, the more there is a retracted zombie trial between authors.

### Retraction cascade following an initial retraction

For the five most prolific first authors in our cohort—Fujii (n=104), Boldt (n=19), Reuben (n=14), Sato (n=11), Saitoh (n=10)—a clear cascade pattern was observed (Figure 4). In each case, an initial retraction was followed by a series of additional retractions involving papers published both before and after the triggering article, sometimes spanning several decades. Fujii represented the most extensive case: a retraction issued in 2012 for a paper published in 2008 trigger paper instantly initiated a massive first wave of synchronous retractions (yellow nodes), prompting a deep historical audit that back-cleared fraudulent publications stretching into the mid-1990s and continuing through 2018. This structural pattern, whereby an initial trigger wave catalyzed the systemic cleanup of an author’s entire publication history, was consistently observed across all five networks.

**Figure 4.**
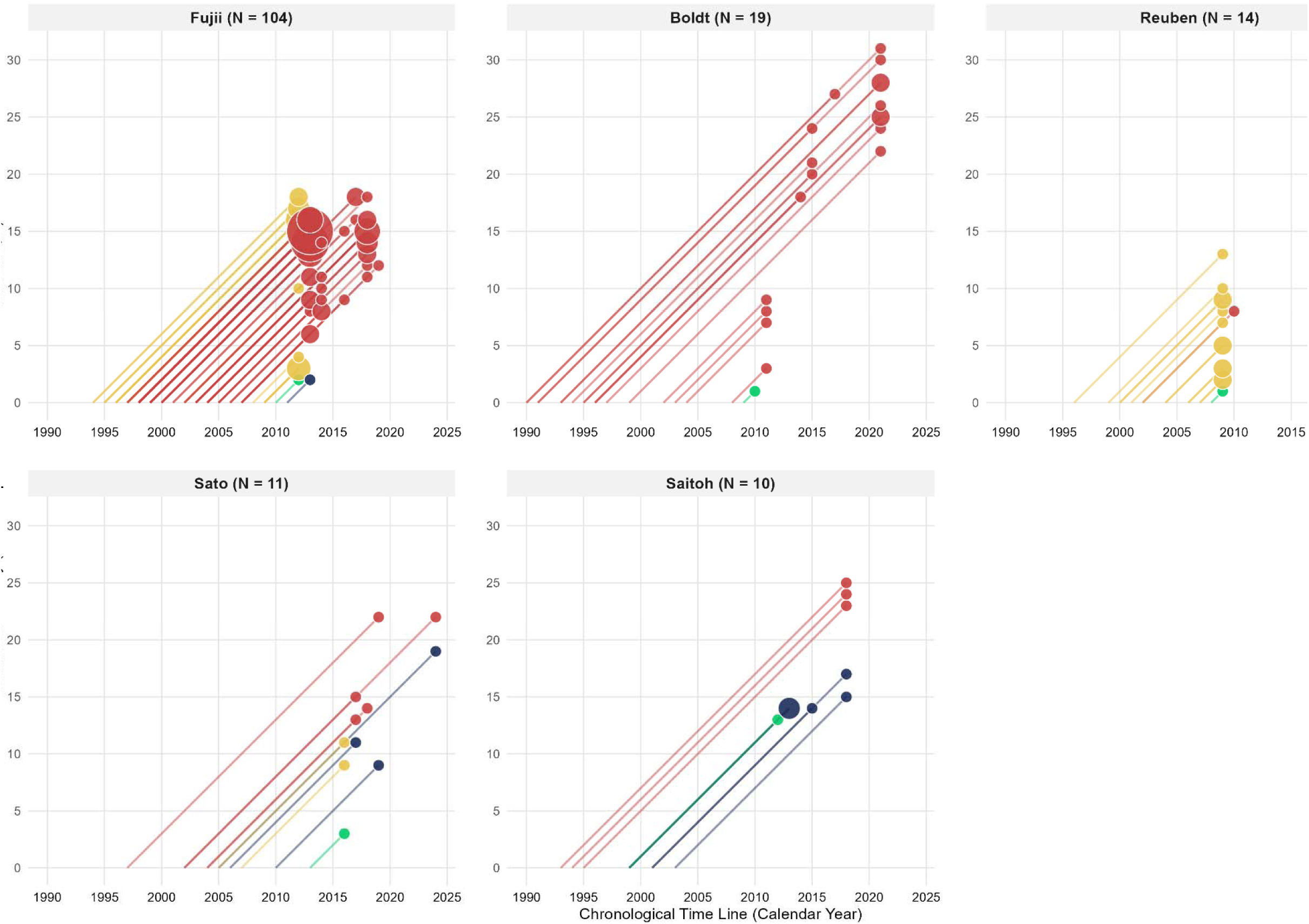
Lexis diagram of retractions relative to the first retraction trigger point. Each panel focuses on one of the top five first authors of retracted zombie trials in our cohort: Fujii (N = 104), Boldt (N = 19), Reuben (N = 14), Sato (N = 11), and Saitoh (N = 10). The horizontal axis tracks chronological calendar time, while the vertical axis indicates retraction latency. Continuous 45◦ diagonal trajectories trace individual papers from their publication year (Y = 0) to their retraction. Terminal event nodes are scaled proportionally to the volume of overlapping timelines sharing identical publication and retraction parameters. Green node Trigger Paper, first retracted zombie trial. Yellow nodes (“First wave”): publications retracted in the same year as the first retracted zombie trial. Red nodes are papers that were older than the first retracted zombie trial, and are retracted after it, and navy-blue nodes are papers that are published after the first retracted zombie trial has been published.

### First and last Author Profile Description

The majority of the 66 (14.7%) first and 100 (22.2%) last authors involved in the publication of retracted zombie trials were male physicians affiliated with public institutions, predominantly specializing in anesthesiology (Appendix VII). Authors associated with multiple publications, whether first or last, exhibited a substantially higher volume of retractions than non-recidivist authors, with a median total of 12.0 retracted trials (IQR 5.0-38.0) compared to 4.5 (IQR 3.0-10.5), respectively.

## Discussion

From an updated cohort of 1,397 retracted RCTs, we identified 236 (16.9%) retracted zombie trials with confirmed data fabrication, of which 197 (83.5%) were linked to authors with a history of repeated misconduct. Those trials were heavily clustered within the field of anesthesiology and the country of Japan. Actually, in the VITALITY cohort, six “superretractors”, as defined by Lyu C et al., co-authored 22% of all retracted trials (24), with several affiliated with Japanese anesthesiology institutions. Notably, three of these individuals (Fujii, Saitoh, and Sato) are represented in our cohort as first authors. These prolific authors appeared to act as central hubs within localized networks of fabricated evidence, sustaining extensive co-authorship networks across fraudulent RCTs. Investigations into these hubs have triggered cascades of retractions spanning decades, with initial retractions exposing earlier fraudulent publications. However, our network analysis revealed fragmented, independent clusters rather than a single, coordinated global web. This implies that fraudulent activity emerged independently across distinct research groups, further supporting the existence of isolated data networks. This is consistent with Fang et al., who showed that a small number of authors account for a disproportionate share of all retractions, and that misconduct underlies 67.4% of all biomedical retractions (25).

Notably, more than four out of five zombie trials were associated with authors who had multiple retractions, suggesting that data fabrication rarely occurs as an isolated event and instead tends to cluster among a limited number of recurrent offenders. Cases with multiple retracted zombie trials tended to be associated with a constellation of characteristics, including lack of trial registration, smaller sample sizes, and absent funding disclosures. These patterns may reflect research environments with reduced transparency and oversight.

Several characteristics of these zombie trials suggest an environment favorable to falsification. Most were monocentric and featured a small author list. One possible explanation is that multicentric studies require greater oversight, multi-institutional collaboration, and independent data management, increasing the likelihood that inconsistencies will be flagged. Similarly, smaller author teams may provide fewer opportunities for independent verification of study conduct (26). Regarding author profiles, the large majority were male physicians working in public institutions specializing in anesthesiology, likely reflecting the baseline demographics of clinical research in these highly affected fields (27). Retracted zombie trials were most often pharmacological in focus and unregistered. The low rate of trial registration is likely explained by the fact that registration was uncommon and not routinely mandated at the time many of these studies were published. However, even among studies published in or after 2005, the registration rate was 27.6%, indicating that trial registration remained incomplete even after the adoption of trial registration policies by medical journals.

In our study, there was an apparent underrepresentation in zombie trials published in recent years. This should not be misconstrued as a reduction in fraud. Given the 12.5-year median retraction delay, recently fabricated trials may simply not have been detected or retracted yet–a structural lag inherent to analyzing retraction timelines. The median time to retraction was particularly prolonged for authors with multiple retractions, reaching 13.6 years (IQR 8.0–15.3). This latency aligns with findings by Nair et al. (27), who reported a median publication-to-retraction delay of 8 years (IQR 3–14). Similarly, Lyu et al. observed that retracted RCTs from “superretractors” had a median delay of 5,111 days (about 14 years) compared to 482 days for other authors (24). Notably, these delays are substantially longer than the general biomedical literature estimate, where the median time from publication to retraction is approximately 2 years (IQR 0.7–4.3) (28).

The observed delays are largely driven by the well-documented case of Yoshitaka Fujii, whose publications account for 104/236 (43.8%) of the cohort. Although concerns regarding the credibility of his findings were first raised in 2000, suspect articles continued to be published and cited for more than a decade before a large-scale retraction process was initiated in 2012 (27). This case illustrates the substantial consequences of delayed retractions: unreliable studies remain part of the evidence base, accumulate citations, and may influence systematic reviews, meta-analyses, and clinical practice guidelines long after concerns have been identified (10,15,29,30).

The concentration of retracted zombie trials in Japan is largely attributable to the Fujii case, though 29 trials (12.3%) from Japan were not first-authored by him. This concentration persisted after adjusting for national clinical trial volume, with Japan retaining the highest ratio among all countries in the cohort (1.8 per 1,000 registered trials). There was also a concentration of retracted zombie trials in anesthesiology, with 137 studies (58.1%), and four of the most prolific retractors (Fujii, Sato, Saitoh, and Reuben) being anesthesiologists. Those four authors account for approximately 59% of all retracted articles in the field (26).

However, the concentration of cases in Japanese anesthesiology should not obscure the phenomenon’s broader scope. Retracted zombie trials were also identified across multiple medical specialties, including cardiology and gynecology, and involved authors from a wide range of countries and institutions. Consistent with our network analysis, the observed patterns suggest that fraudulent RCTs emerged through multiple independent research networks rather than a single disciplinary or geographical setting. Moreover, these studies were published in a diverse range of journals and by various publishers, highlighting that the persistence of zombie trials reflects not only individual misconduct but also vulnerabilities within the broader scientific publication ecosystem. Finally, countries and fields with higher rates of zombie retracted trials may not necessarily have higher rates of misconduct; they may simply be more sensitive and rigorous and thus likely to detect and act upon misconduct.

## Limitations

Given the descriptive nature of this study, apparent geographical, specialty-specific, or publisher-specific clustering of zombie trials should be interpreted cautiously and not considered evidence of underlying causal mechanisms.

In prominent cases of repeated misconduct, retraction notices may be more likely to explicitly identify data fabrication, whereas fabrication involving other authors may be described ambiguously or not mentioned at all. Consequently, our cohort does not capture all fabricated studies, and the apparent concentration of zombie trials across countries, specialties, publishers, journal impact factors, or possible predatory status may partly reflect differences in the transparency of retraction practices, post-publication scrutiny, proactive screening, and research integrity policies rather than the true distribution of research misconduct. Without transparency regarding the number of investigations conducted by publishers, these explanations cannot be disentangled.

Importantly, given the median retraction delay of 13.6 years, many fabricated trials published over the last decade have likely not yet been identified or retracted. In addition, some studies containing fabricated data may remain undetected, and even when fabrication is identified, editorial action is not always taken. Finally, our identification strategy relied on text-mining retraction notices for specific terms, so cases described using vague, euphemistic, or non-standard language may have been missed. Consequently, the true prevalence of zombie trials cannot be inferred from this sample; rather, we established a cohort of trials with clear evidence of fabrication, recognizing that retraction notices are often incomplete regarding the reasons for retraction.

## Conclusion

Zombie trials pose a persistent threat to scientific integrity. This cohort of retracted zombie trials helps characterize who produces them, what features they share, and how they propagate through scientific literature. Identifying indicators that may facilitate earlier detection of zombie trials, limit their dissemination, and safeguard the integrity of the evidence base is highly desirable. Emerging detection tools, such as INSPECT-SR (33), represent a further step in this effort. Since INSPECT-SR is not designed to confirm data fabrication but rather to flag trials with signals of untrustworthiness, this cohort of confirmed zombie trials offers a valuable ground-truth resource to assess whether such tools correctly characterize these trials as problematic.

## Data Availability

All data produced in the present study are available upon reasonable request to the authors

## Acknowledgements

We would like to thank Xu et al. for making their cohort openly accessible, which served as the foundation for our dataset.

## Authors’ contributions

Tala Jajieh: Writing—Original draft, Writing—review and editing, Visualization, Software, Methodology, Formal analysis, Data curation. Silvy Laporte: Validation, Supervision, Project administration, Methodology, Conceptualization, Writing—review and editing. Florian Naudet: Conceptualization, Methodology, Funding acquisition, Writing—review and editing. Céline Chapelle: Validation, Methodology, Writing—review and editing. Edouard Ollier: Writing— review and editing. Marc-Antoine Pencolé: Writing—review and editing. Cédric Lemarchand: Writing—review and editing. Clara Locher: Writing—review and editing. John P.A. Ioannidis: Methodology, Writing—review and editing.

## AI contribution disclosure

The author declares the use of generative artificial intelligence (GAI) in the writing process. According to the GAIDeT taxonomy, the following tasks were delegated to GAI tools under full human supervision: proofreading and editing. The GAI tool used was ChatGPT-5.5 (OpenAI). Responsibility for the content and the final manuscript lies entirely with the author. After using this tool, the author thoroughly reviewed, revised, and edited the manuscript, and takes full responsibility for its final version.

## References

1. Agence nationale de la recherche [Internet]. [cited 2025 Oct 14]. Scientific Integrity. Available from: https://anr.fr/en/anrs-role-in-research/commitments/scientific-integrity/

2. EurekAlert! [Internet]. [cited 2025 Oct 8]. Organized scientific fraud is growing at an alarming rate. Available from: https://www.eurekalert.org/news-releases/1093143

3. Definition of Research Misconduct | ORI - The Office of Research Integrity [Internet]. [cited 2025 Oct 8]. Available from: https://ori.hhs.gov/definition-research-misconduct

4. Retraction Watch [Internet]. 2026 [cited 2026 Jul 2]. June 2026. Available from: https://retractionwatch.com/2026/06/

5. Van Noorden R. More than 10,000 research papers were retracted in 2023 — a new record. Nature. 2023 Dec 12;624(7992):479–81. doi:10.1038/d41586-023-03974-8

6. Loadsman JA. Why does retraction take so much longer than publication? Anaesthesia. 2019;74(1):3–5. doi:10.1111/anae.14484

7. Sibbald B, Roland M. Understanding controlled trials: Why are randomised controlled trials important? BMJ. 1998 Jan 17;316(7126):201. doi:10.1136/bmj.316.7126.201 PubMed PMID: 9468688.

8. Users’ Guides to the Medical Literature: A Manual for Evidence-Based Clinical Practice, 3rd ed | JAMAevidence | McGraw Hill Medical [Internet]. [cited 2025 Mar 5]. Available from: https://jamaevidence.mhmedical.com/book.aspx?bookId=847

9. Graña Possamai C, Cabanac G, Perrodeau E, Ghosn L, Ravaud P, Boutron I. Inclusion of Retracted Studies in Systematic Reviews and Meta-Analyses of Interventions: A Systematic Review and Meta-Analysis. JAMA Intern Med. 2025 Mar 31;e250256. doi:10.1001/jamainternmed.2025.0256 PubMed PMID: 40163084; PubMed Central PMCID: PMC11959482.

10. Investigating the impact of trial retractions on the healthcare evidence ecosystem (VITALITY Study I): retrospective cohort study | The BMJ [Internet]. [cited 2025 Apr 30]. Available from: https://www.bmj.com/content/389/bmj-2024-082068

11. Van Noorden R. Medicine is plagued by untrustworthy clinical trials. How many studies are faked or flawed? Nature. 2023 Jul 18;619(7970):454–8. doi:10.1038/d41586-023-02299-w

12. Else H. What universities can learn from one of science’s biggest frauds. Nature. 2019 Jun 18;570(7761):287–8. doi:10.1038/d41586-019-01884-2

13. Avenell A, Bolland MJ, Gamble GD, Grey A. A randomized trial alerting authors, with or without coauthors or editors, that research they cited in systematic reviews and guidelines has been retracted. Account Res. 2024 Jan 2;31(1):14–37. doi:10.1080/08989621.2022.2082290

14. Ioannidis JPA. Hundreds of thousands of zombie randomised trials circulate among us. Anaesthesia. 2021 Apr;76(4):444–7. doi:10.1111/anae.15297

15. Carlisle JB. False individual patient data and zombie randomised controlled trials submitted to *Anaesthesia*. Anaesthesia. 2021 Apr;76(4):472–9. doi:10.1111/anae.15263

16. Mol BW, Ioannidis JPA. How do we increase the trustworthiness of medical publications? Fertil Steril. 2023 Sep;120(3):412–4. doi:10.1016/j.fertnstert.2023.02.023

17. Developing a tool for detecting problematic RCTs in health systematic reviews: the INSPECT-SR project [Internet]. [cited 2025 Mar 6]. Available from: https://training.cochrane.org/resource/msu-web-clinic-july-2023

18. Page MJ, McKenzie JE, Bossuyt PM, Boutron I, Hoffmann TC, Mulrow CD, et al. The PRISMA 2020 statement: an updated guideline for reporting systematic reviews [Internet]. 2021 Mar 29. doi:10.1136/bmj.n71

19. PRISMA statement [Internet]. [cited 2026 Jun 18]. PRISMA 2020 flow diagram. Available from: https://www.prisma-statement.org/prisma-2020-flow-diagram

20. Retraction Watch Database [Internet]. [cited 2025 Mar 6]. Available from: https://retractiondatabase.org/RetractionSearch.aspx?

21. PubPeer - Search publications and join the conversation. [Internet]. [cited 2025 Mar 6]. Available from: https://pubpeer.com/static/about

22. Rittman M. Crossref [website] [Internet]. [cited 2026 Jan 7]. REST API. Available from: https://www.crossref.org/documentation/retrieve-metadata/rest-api/

23. Number of clinical trials by year, country, region and income group [Internet]. [cited 2026 Jul 7]. Available from: https://www.who.int/observatories/global-observatory-on-health-research-and-development/monitoring/number-of-clinical-trials-by-year-country-who-region-and-income-group

24. Lyu C, Matbouriahi M, Naudet F, Ioannidis JPA, Cristea IA. Retracted Randomized Clinical Trials From Superretractors and Top-Cited Scientists With Multiple Retractions. JAMA Netw Open. 2026 Apr 1;9(4):e267424. doi:10.1001/jamanetworkopen.2026.7424 PubMed PMID: 41984475; PubMed Central PMCID: PMC13084457.

25. Fang FC, Steen RG, Casadevall A. Misconduct accounts for the majority of retracted scientific publications. Proc Natl Acad Sci U S A. 2012 Oct 16;109(42):17028–33. doi:10.1073/pnas.1212247109 PubMed PMID: 23027971; PubMed Central PMCID: PMC3479492.

26. Mongeon P, Larivière V. Costly collaborations: The impact of scientific fraud on co authors’ careers. J Assoc Inf Sci Technol. 2015. doi:10.1002/asi.23421

27. Nair S, Yean C, Yoo J, Leff J, Delphin E, Adams DC. Reasons for article retraction in anesthesiology: a comprehensive analysis. Can J Anaesth J Can Anesth. 2020 Jan;67(1):57–63. doi:10.1007/s12630-019-01508-3 PubMed PMID: 31617069.

28. Ferraro MC, Moore RA, de C Williams AC, Fisher E, Stewart G, Ferguson MC, et al. Characteristics of retracted publications related to pain research: a systematic review. Pain. 2023 Nov 1;164(11):2397–404. doi:10.1097/j.pain.0000000000002947 PubMed PMID: 37310441.

29. Weibel S, Popp M, Reis S, Skoetz N, Garner P, Sydenham E. Identifying and managing problematic trials: A research integrity assessment tool for randomized controlled trials in evidence synthesis. Res Synth Methods. 2023 May;14(3):357–69. doi:10.1002/jrsm.1599 PubMed PMID: 36054583; PubMed Central PMCID: PMC10551123.

30. Kataoka Y, Banno M, Tsujimoto Y, Ariie T, Taito S, Suzuki T, et al. Retracted randomized controlled trials were cited and not corrected in systematic reviews and clinical practice guidelines. J Clin Epidemiol. 2022 Oct;150:90–7. doi:10.1016/j.jclinepi.2022.06.015 PubMed PMID: 35779825.

31. A systematic review of retractions in biomedical research publications: reasons for retractions and their citations in Indian affiliations | Humanities and Social Sciences Communications [Internet]. [cited 2026 Jun 3]. Available from: https://www.nature.com/articles/s41599-023-02095-x

32. Kocyigit BF, Akyol A. Analysis of Retracted Publications in The Biomedical Literature from Turkey. J Korean Med Sci. 2022 May 9;37(18):e142. doi:10.3346/jkms.2022.37.e142 PubMed PMID: 35535370; PubMed Central PMCID: PMC9091427.

33. Wilkinson J, Heal C, Flemyng E, Antoniou GA, Aburrow T, Alfirevic Z, et al. INSPECT-SR: a tool for assessing trustworthiness of randomised controlled trials. medRxiv: The Preprint Server for Health Sciences; 2025. doi:10.1101/2025.09.03.25334905 PubMed PMID: 40950444; PubMed Central PMCID: PMC12424918.

34. RestoRes | Research integrity in biomedical research [Internet]. [cited 2025 Mar 5]. Available from: https://restores.univ-rennes.fr/

