## Supplementary Material for "Zombie trials are often monocentric and cluster within fabricated evidence “factories”: a cohort study of 236 retracted randomized controlled trials with confirmed data fabrication"

**Affiliations**

Tala Jajieh 0009-0009-7459-5426

Céline Chapelle 0000-0003-3281-3666

Cédric Lemarchand 0009-0009-8695-1248

Clara Locher 0000-0002-8212-4351

Marc-Antoine Pencolé 0000-0002-3497-7966

Edouard Ollier 0000-0001-6925-2941

John P.A. Ioannidis 0000-0003-3118-6859

Florian Naudet 0000-0003-3760-3801

Silvy Laporte 0000-0001-6197-8668

### Appendix I. Text-Mining Search Strategy for Identification of Retracted Zombie Trials

*Keywords and data sources used to identify retracted zombie trials within the updated VITALITY cohort.*

| **Step** | **Description** |
| --- | --- |
| **Data source** | Updated (through 11 December 2025) Retraction Watch database — VITALITY cohort of 1,397 retracted RCTs |
| **Text-mining keywords** | not authentic; authenticity; fictive; fictitious; fabrication; fabricated; fabricate; imagination |
| **Fields searched** | Reason of retraction AND retraction notice AND/OR PubPeer post-publication peer review comments |
| **Method** | Text mining performed using R software |
| **Result** | 236 retracted zombie trials identified |

### Appendix II. Data Extraction Variables and Sources

*Complete list of variables extracted for each retracted zombie trial, including extraction method and data source.*

| **Data** | **Extraction Type** | **Source** |
| --- | --- | --- |
| DOI | Automatic | Retraction Watch |
| PMID | Automatic | Retraction Watch |
| Title | Automatic | Retraction Watch |
| Authors | Automatic | Retraction Watch |
| First author / last author | Generated | — |
| Author's recidivism status | Generated | — |
| Number of authors in each paper | Generated | — |
| Journal | Automatic | Retraction Watch |
| Recommendable journal* | Generated | — |
| Year of publication | Automatic | Retraction Watch |
| Retraction date | Automatic | Retraction Watch |
| Reason for retraction | Automatic | Retraction Watch |
| Author approval for retraction | Manual | Reason for retraction |
| Country | Automatic | Retraction Watch |
| Continent | Automatic | Country variable |
| Medical specialty | Automatic (and manual if needed) | Retraction Watch |
| Publishers | Automatic | Retraction Watch |
| Paywalled | Automatic | Retraction Watch |
| Submission date | Automatic | PubMed |
| Acceptance date | Automatic | PubMed |
| Retraction notice | Manual | PubMed |
| Registration | Manual | ClinicalTrials.gov and open-access paper |
| Registration number / Funding | Manual | ClinicalTrials.gov and open-access paper |
| Sample size | Manual | Open-access paper |
| Intervention | Manual | Open-access paper |
| Intervention class | Generated | Intervention variable |
| Primary outcome significance (p-value) | Manual | Open-access paper |
| Journal Impact Factor | Manual | Web of Science |
| Retraction delay | Generated | — |
| Author variables: sex / institutional affiliation / clinical specialty / academic credentials (MD vs non-MD status) | Manual | ResearchGate, LinkedIn, PubMed, Google Scholar |

**Recommendable journal (non-predatory) status was assessed using the list of recommended journals published by the Conférence des Doyens de Médecine and the CNU Santé (conferencedesdoyensdemedecine.org), which lists journals presumed not to be predatory across health, medicine, and biology fields. (the list date November 2025)*

### Appendix III. Multiple Correspondence Analysis of retracted zombie trial characteristics

**(A)** Symmetric plot of variables (contributing characteristics). This variable map gives a visual picture of how the characteristics of retracted zombie trials relate to one another. Dimension 1 (horizontal axis) and dimension 2 (vertical axis) explain 18.1% and 12.7%, respectively, accounting for 30.8% combined. The further a modality sits from the center of the plot, the more it contributes to distinguishing between author profiles. **(B)** MCA biplot. Each dot represents a retracted zombie trial; the darker the dot reflects more trials share similar profiles. The 95% confidence ellipses around each group help visualize how distinct the two profiles are.

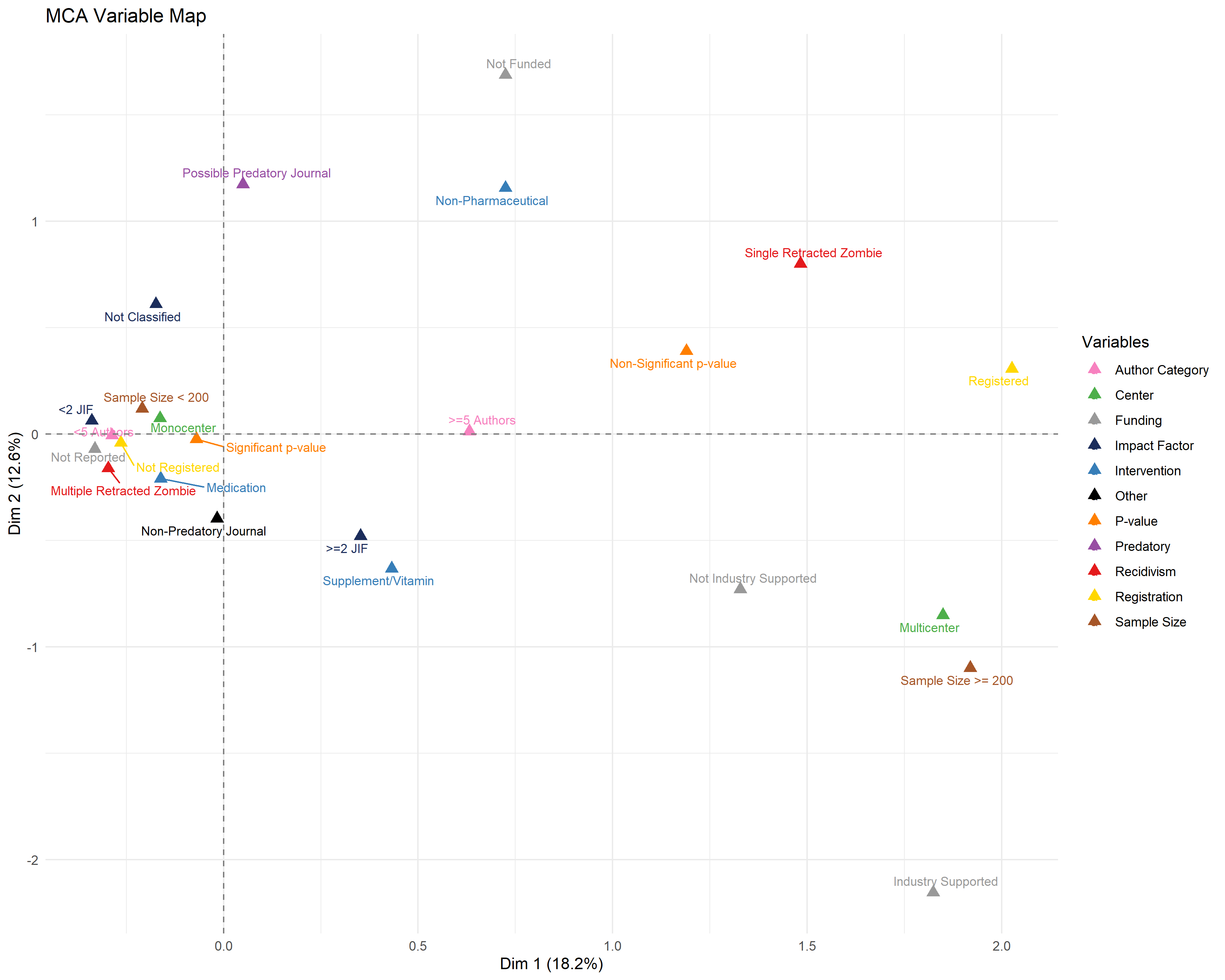
**A.**

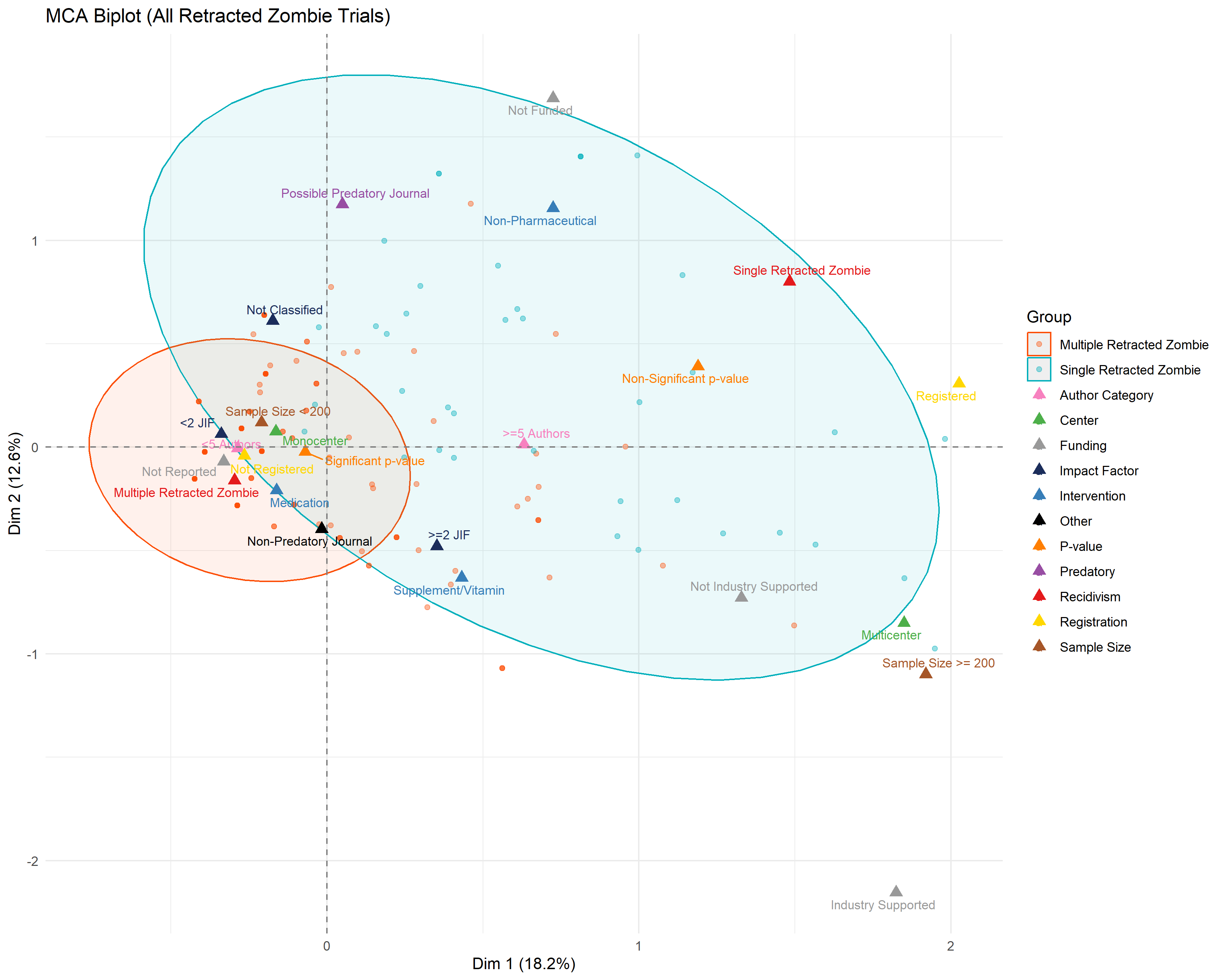
**B**

### Appendix IV. Shift in country ranking of retracted zombie trials after adjustment for national clinical trial volume, using fractional country attribution.

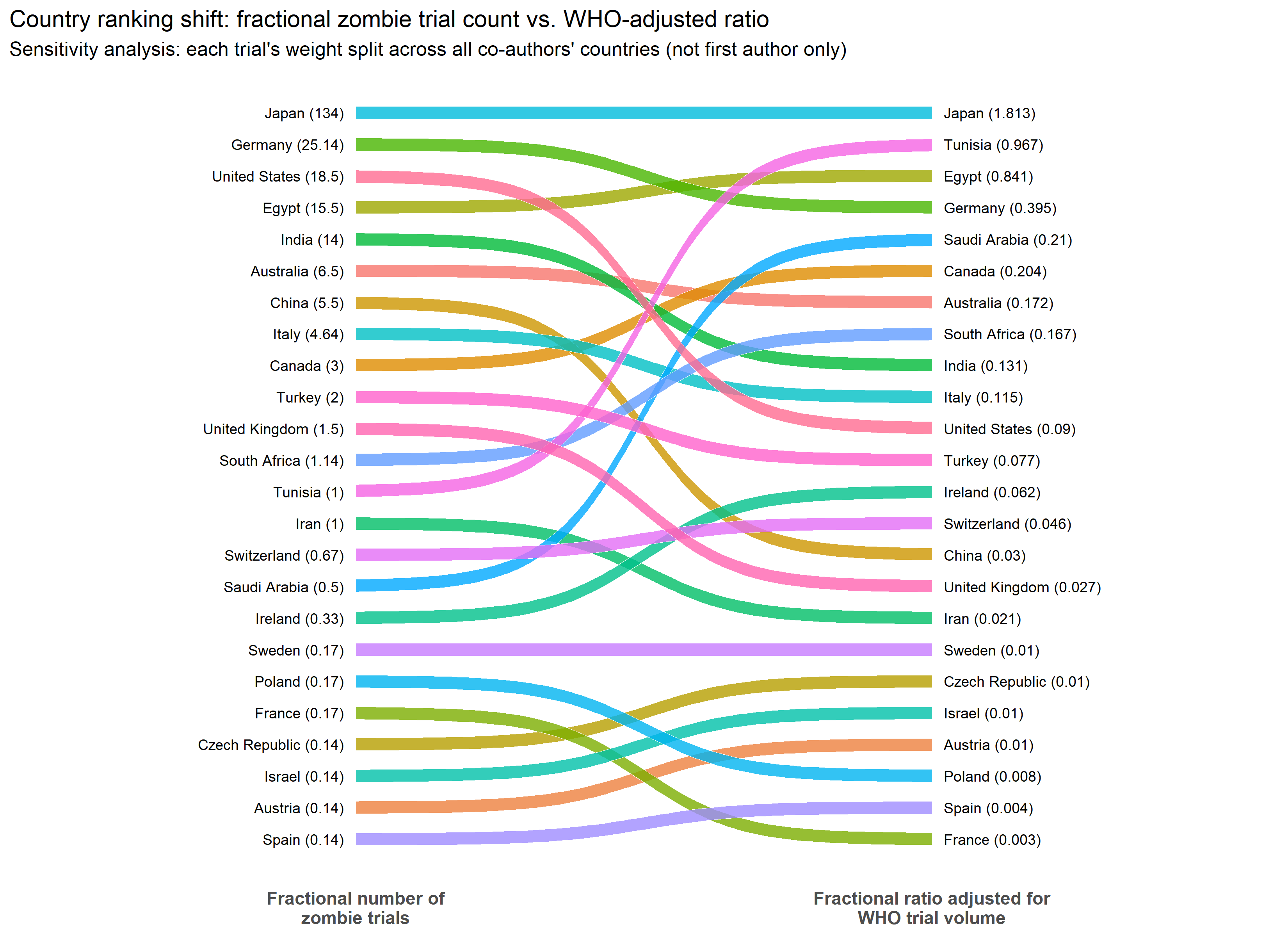
Comparison of country ranking by fractional number of retracted zombie trials (left, per 236 total zombie trials) versus ranking adjusted for each country's total registered clinical trial volume (right, per 1,000 registered clinical trials). Fractional counts represent each trial's contribution divided equally among all countries represented among its authors, rather than the first author's country alone. Adjusted ratios represent the fractional number of retracted zombie trials per 1,000 clinical trials registered in that country, using trial volume data from the WHO Global Observatory on Health Research and Development. Colored bands connect each country's position across the two rankings; countries are ordered from highest to lowest within each column.

### Appendix V. Geographical Distribution of Retracted Zombie Trials and All Contributing Authors

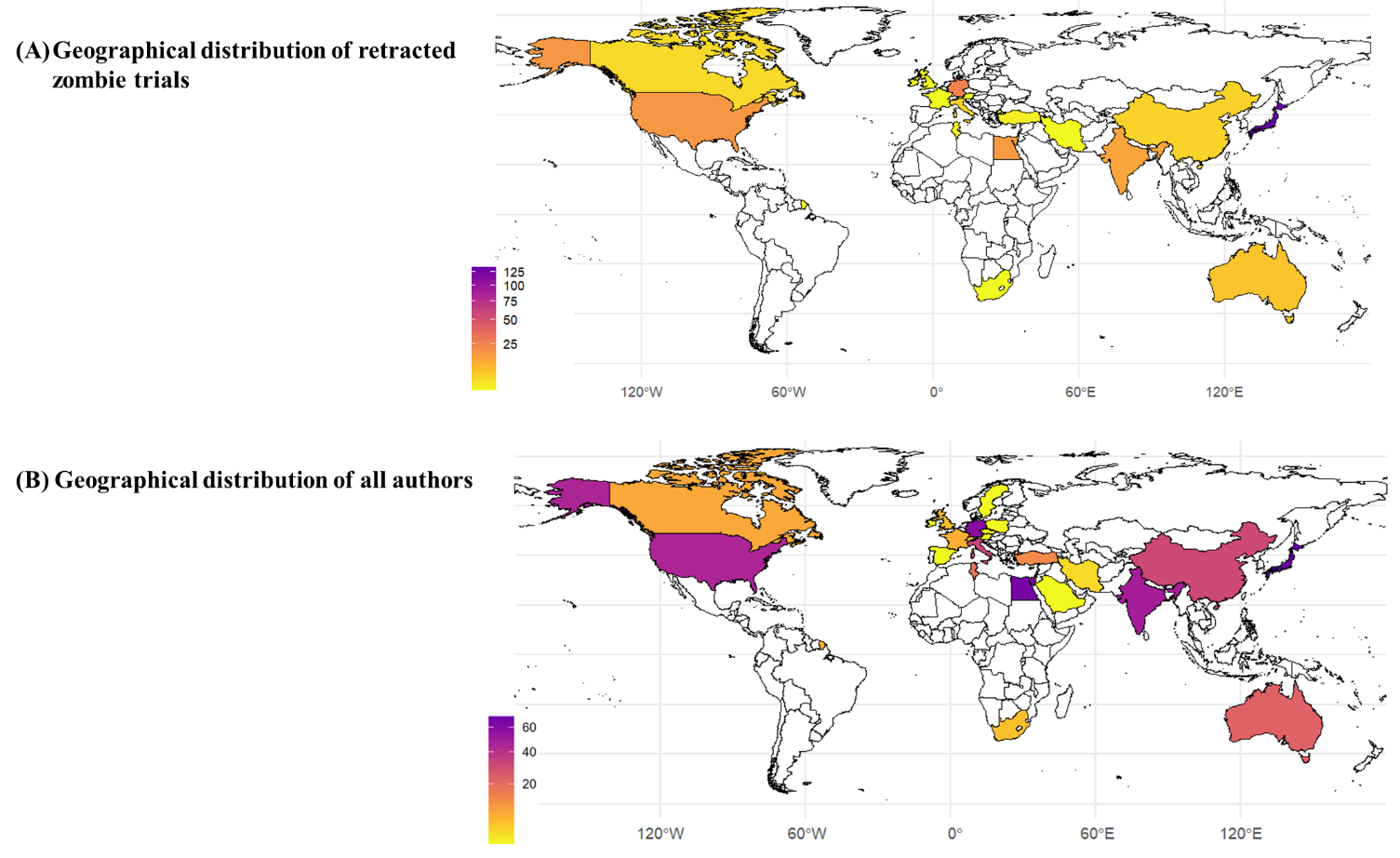
(A) Geographical Distribution of Retracted Zombie Trials without adjustment for national clinical trial volume. (B)Geographical distribution of all Authors of retracted zombie trials. Color intensity reflects the number of retracted Zombie Trials(A) and of authors (B).

**A.**

**B.**

### Appendix VI. Comparison of PubMed-Indexed Articles versus Retracted Zombie Trials by Publisher

*Distribution of retracted zombie trials across publishers (orange) relative to their overall publication volume indexed in PubMed (blue) .*

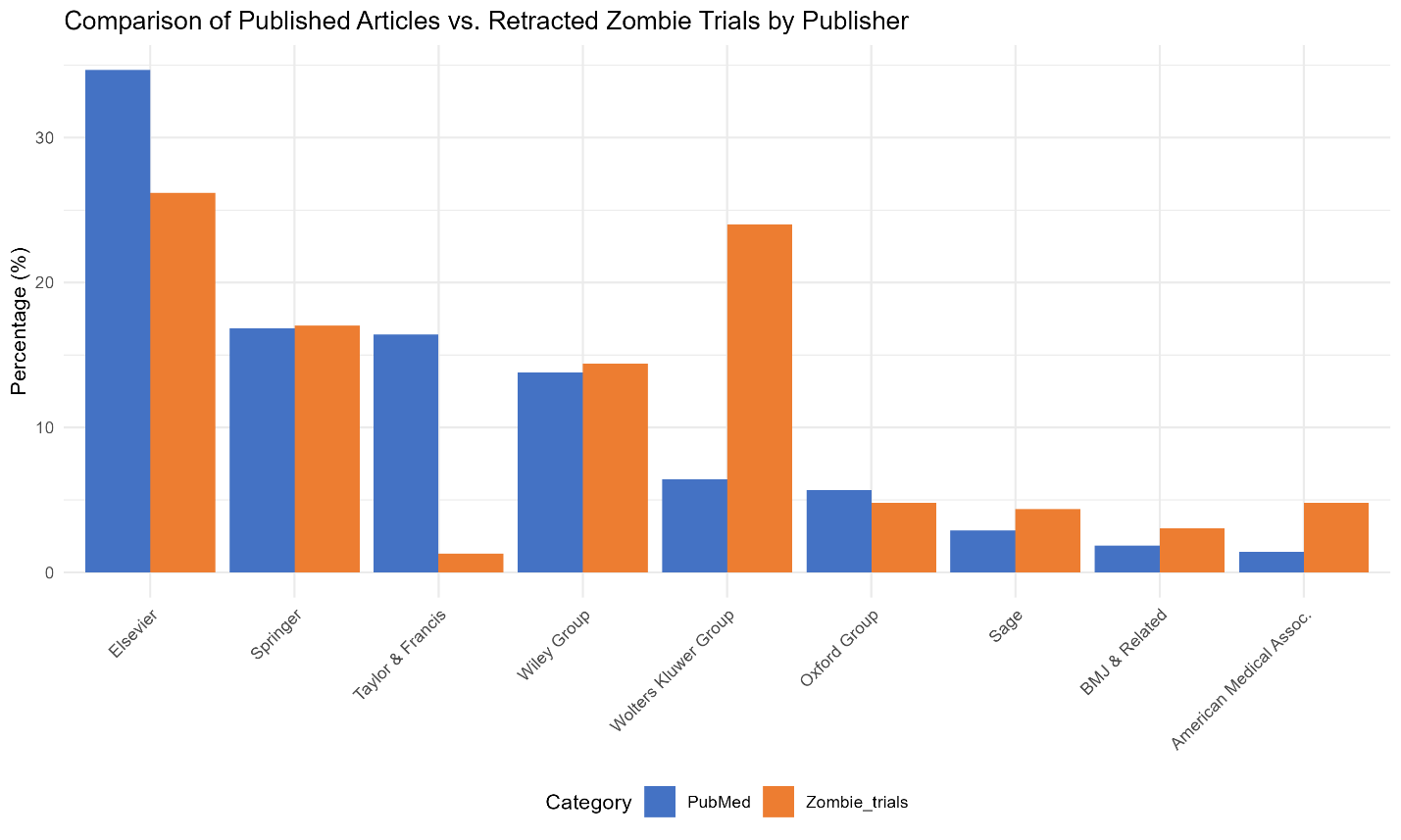

### Appendix VII. Comparison of First and Last Authors' Profiles Associated with Multiple Versus Single Retracted Zombie Trials

*Demographic, professional, and retraction-history characteristics of first and last authors, stratified by single versus multiple retracted zombie trial involvement.*

|  | **First Author (N = 66)** | | **Last Author (N = 100)** | |
| --- | --- | --- | --- | --- |
| **Variable** | **Single (N=48)** | **Multiple (N=18)** | **Single (N=59)** | **Multiple (N=41)** |
| Male gender, n (%) | 43 (89.6) | 17 (94.4) | 49 (83.1) | 34 (82.9) |
| Physician, n (%) | 34 (70.8) | 15 (83.3) | 46 (78.0) | 29 (70.7) |
| Public-affiliated institution, n (%) | 41 (85.4) | 13 (72.2) | 56 (94.9) | 38 (92.7) |
| **Specialty, n (%)** | | | | |
| Anesthesiology | 18 (37.5) | 10 (55.5) | 16 (27.1) | 28 (68.3) |
| Gynaecology | 8 (16.7) | 3 (16.7) | 7 (11.9) | 2 (4.9) |
| Oncology | 4 (8.3) | 0 (0) | 3 (5.1) | 1 (2.4) |
| Others | 18 (37.5) | 5 (27.8) | 33 (55.9) | 10 (24.4) |
| **Retraction history, median (IQR)** | | | | |
| Total retracted trials | 1.0 (1.0–3.0) | 12.0 (5.0–38.0) | 1.0 (1.0–1.0) | 4.5 (3.0–10.5) |
| Zombie retracted trials | 1.0 (1.0–1.0) | 11 (3.5–17.5) | 1.0 (1.0–1.0) | 3 (2.0–6.0) |
| Retracted non-zombie | 0 (0–1.0) | 2.0 (1.0–11.0) | 0 (0–0) | 1.5 (0–5.0) |

### Appendix VIII. R Packages Used for Statistical Analysis and Data Visualization

*R packages used at each stage of data extraction, cleaning, statistical analysis, and visualization.*

| **Package** | **Used for** |
| --- | --- |
| **PRISMA flowchart and study selection** | |
| *dplyr* | Data filtering, deduplication, and selection logic for the PRISMA flow |
| **RCT verification (PubMed cross-check)** | |
| *rentrez* | Querying PubMed via the Entrez API to verify publication type |
| *dplyr* | Data wrangling of verification results |
| **Text-mining (zombie trial detection)** | |
| *dplyr* | Keyword pattern matching and classification of retraction reasons/notes (base R grepl() used for the pattern search itself) |
| *stringi* | Encoding cleanup of text fields prior to text-mining (stri_replace_all_fixed, stri_enc_toutf8) |
| **Author-level recidivism classification (first and last authors)** | |
| *dplyr* | Author-level grouping, counting, and recidivism status joins |
| *stringr* | String normalization (str_to_lower, str_trim) |
| *readr* | CSV import/export (write_csv) |
| *tidyr* | Splitting multi-author strings into individual rows (separate_rows) |
| *ggplot2* | Bar plots of top contributing authors |
| **Co-authorship network analysis** | |
| *igraph* | Graph construction, simplification, and component/cluster detection |
| *ggraph* | Network visualization (edges, nodes, layouts) |
| *dplyr* | Edge list and node summary construction |
| *tidyr* | Reshaping author-pair combinations (unnest()) |
| *ggplot2* | Plot theming and labels |
| *ggrepel* | Non-overlapping node labels in multi-component plots |
| **Multiple Correspondence Analysis (MCA)** | |
| *FactoMineR* | MCA computation |
| *factoextra* | MCA visualization helpers (fviz_mca_var, fviz_mca_biplot) |
| *dplyr* | Variable recoding and data preparation |
| *tibble* | Data frame handling |
| *ggplot2* | Custom biplot and variable map construction |
| *ggrepel* | Non-overlapping variable labels |
| **Lexis diagram (retraction cascade analysis)** | |
| *dplyr* | Data filtering, grouping, and cascade metric calculation |
| *ggplot2* | Diagram rendering (segments, points, facets) |
| *tibble* | Data frame handling |
| **Geographic distribution mapping** | |
| *dplyr* | Country-level aggregation |
| *countrycode* | Country name standardization |
| *ggplot2* | Choropleth map rendering |
| *rnaturalearth / rnaturalearthdata* | World map shapefiles |
| *viridis* | Color scale for choropleth gradients |
| *sf* | Spatial data handling |
| *stringr* | String splitting/trimming for multi-author/multi-country fields |
| *tidyr* | Unnesting author-country pairs (unnest_longer()) |
| **Publisher-level PubMed benchmarking (journal-to-publisher mapping)** | |
| *httr* | Querying the Crossref REST API for journal/publisher metadata |
| *jsonlite* | Parsing JSON responses from the Crossref API |
| *dplyr* | Filtering and grouping journals by publisher |
| *rentrez* | Querying PubMed for per-journal article counts to estimate publisher-level baseline publication volume |
